# Developing and testing Advance Choice Document implementation resources for Black African and Caribbean people with experience of compulsory psychiatric admission

**DOI:** 10.1101/2024.04.17.24305837

**Authors:** Jonathan Simpson, Abigail Babatunde, Alan Simpson, Steven Gilbert, Alex Ruck Keene, Lucy Stephenson, Kia-Chong Chua, Gareth Owen, Fiona Crowe, Pauline Edwards, Selena Galloway, Megan Fisher, Marcela Schilderman, Anita Bignell, Shubulade Smith, Claire Henderson

## Abstract

**Background:** Advance Choice Documents (ACDs) have been recommended for use in England and Wales based on evidence from trials that show that they can reduce involuntary hospitalisation, which disproportionately affects Black African and Caribbean people. Our aim was therefore to develop and test ACD implementation resources and processes for Black people who have previously been involuntarily hospitalised and the people that support them.

**Methods:** Resource co-production workshops were held to inform the development of the ACD template and two types of training for all stakeholders, comprising a Recovery College course and simulation training. An ACD facilitator then used the ACD template developed through the workshops to create personalised ACDs with service users and mental health staff over a series of meetings. Interviews were then conducted with service user and staff participants and analysed to document their experience of the process and opinions on ACDs. Other implementation strategies were also employed alongside to support and optimise the creation of ACDs.

**Results:** Nine ACDs were completed and were largely reported as appropriate, acceptable, and feasible to service users and staff. Both reported it being an empowering process that encouraged hope for better future treatment and therefore better wellbeing. Uncertainty was also expressed about the confidence people had that ACDs would be adhered to/honoured, primarily due to staff workload. The information provision training and the skills training were generally considered to be informative by trainees.

**Conclusions:** The project has developed an ACD creation resource that was reported as agreeable to all stakeholders; however, the generalisability of the findings is limited due to the small sample size. The project also highlights the importance of staff and ACD facilitator capacity and good therapeutic relationships in ACD completion. Further research is needed to determine the adjustments needed for large scale use, including for those under age 18 and those under the care of forensic mental health services; and how to include carers/supporters more in the process.

## Background

It is consistently found that Black people, defined as people of Black African and Caribbean heritage and people of mixed Black heritage, disproportionally have negative experiences of mental health services in the UK (1–3). This includes more coercive treatment, higher police involvement in detention in psychiatric hospitals under the Mental Health Act (MHA), and more frequent and sustained compulsory detentions (1,4) which all act to reduce autonomy. These disparities also exist alongside poorer mental health outcomes (2,5). Interventions designed to improve the experience of Black people in mental health services have been insufficient (6). However, written statements of treatment preferences created in advance of a mental health crisis or relapse have been associated with reduced compulsory psychiatric admissions (7–9) and reduced periods as an inpatient for Black people in particular (10). These are referred to as Advance Choice Documents (ACDs) in the 2018 Independent Review of the UK Mental Health Act (MHA Review) and depending on the legislative context and authors are variously termed Advance Statements, Psychiatric Advance Directives, Joint Crisis Plans, and Crisis Cards.

ACDs can support the autonomy of service users (6,11,12) and facilitate mental health care providers being held accountable in delivering preferred care. They can potentially address Black service users’ dissatisfaction with services (13), impaired therapeutic alliance and trust, and reduced help seeking (14,15). ACDs are viewed positively by service users, carers/supporters of people with mental health difficulties, and mental health staff (MHS) (16,17). The UK government accepted the recommendation of the use of ACDs in the MHA Review and the parliamentary scrutiny committee (which provides specialist expertise on draft bills as part of the Committee Office in the House of Commons) recommended a statutory offer be made to anyone who has previously been detained under the MHA (18). However, in countries where there is legislation supporting the use of ACDs, such as Scotland and the USA, uptake is low (19).

Various reasons have been cited for the lack of creation and use of ACDs, at the level of the service user, staff, and service (20). A study of Joint Crisis Plan use identified three barriers: 1) lack of recognition of the benefits of ACDs; 2) not recognising the need for a change in the clinician-service user relationship, including discussing treatment options and supporting patient choice; and 3) difficulties in implementation when working across the healthcare system (17).

At the time of writing, legislation is still pending; however, ACD implementation has been incorporated by NHS England into both its Patient and Carer Race Equality Framework reporting requirements (21) and its recent guidance ‘Acute inpatient mental health care for adults and older adults’(22). ACD creation and use is therefore currently seen as good practice. To support effective implementation of ACDs by National Health Service (NHS) mental health service providers and for wider adoption, this implementation resource development and testing research project (the Advance Statements for Black African and Caribbean people project; AdStAC) consisted of three phases: stakeholder workshops (to understand views about ACDs) (23); co-production (deciding what resources were needed to implement ACDs and in what format); and implementation (testing and revising the implementation resource) (24). This paper focuses on the co-production and implementation phases, which had the aims:

1. Co-produce an implementation resource (guidance and training on ACD implementation, and a modified ACD template)
2. Test, revise, and re-test the ACD implementation resources developed

## Methods

### Design

The resource co-production phase comprised three workshops: ACD template design; Recovery College training on understanding ACDs, and simulation training on how to create and use ACDs.

The implementation phase comprised Plan-Do-Study-Act cycles (25) to create ACDs, gain feedback, and modify the processes as necessary.

### Ethics

Approval for the study was granted by the Bradford Leeds NHS Health Research Authority Research Ethics Committee (Research Ethics Committee reference number: 22/YH/0012) on 07/02/2022.

### Context

The sites where the implementation resources were tested were in four South London local government areas served by an NHS mental health service provider (an NHS Trust) that have, relative to the UK, a high proportion of Black people (range 22.6%-26.8%) (26).

The NHS Trust has a Recovery College, which offers courses and workshops taught by people with lived experience of long-term mental health conditions/difficulties and people with expertise by profession. Recovery Colleges are widespread throughout the UK (88 in 2021; (27) and are open to service users, carers/supporters, and MHS to attend. The Trust also has a mental health simulation training centre that offers courses that can be booked by any NHS Trust.

### Participants

The workshops were open to Black people aged 16+ who had previously been detained under the MHA; supporters/carers of a Black service user eligible for the project; MHS who may be involved in supporting the completion of ACDs/refer to ACDs in clinical records/involved in compulsory admission, and General Practitioners as they are likely to work with people who have severe mental illness.

The targeted groups for the intervention (both sets of training and ACD creation) were Black service users and the people who support and work with them. Inclusion criteria for service users included: currently using the NHS Trust services; aged 16+; previously detained at least once under the MHA; and self-identifying as of Black African and Caribbean heritage, or of mixed Black heritage. For supporters/carers: aged 16+; informal carers/supporters of eligible service users; and likely to be named in the ACD. Eligible staff included those who may be involved in completing or referring to the ACD in clinical records, such as MHS employed by the Trust in acute and community settings, section 12 (MHA) approved doctors, Approved Mental Health Professionals, advocates, and peer workers.

Purposive sampling was used to recruit service users, carers/supporters, and staff for the co-production workshops and the trainings; separate recruitment campaigns were carried out for each sets of training and the ACD creation. Recruitment occurred through presentations to clinical teams and third-sector organisations (including community groups, voluntary organisations, and faith and equalities groups). Physical and digital adverts were also distributed to these groups and promoted via social media platforms. For the creation of ACDs, the first 15 eligible service users expressing interest were recruited through referrals from MHS and third-sector organisation staff, as well as via the Trust’s consent for contact register. Initially the delivery of sixty ACDs was planned to have information power for several Plan Do Study Act cycles (25); however, it was only possible to employ a facilitator part-time and thus we were unable to achieve this sample size within the study period.

### Stakeholder workshops guidance

The stakeholder workshops phase of the project provided guidance around the approach to developing and testing the ACD implementation resource (23). The workshops with service users, carers/supporters, and MHS highlighted key themes of ‘people’ ‘process’ and ‘power’ as important when considering ACD implementation.

A suggestion from the workshops was to include a section in the ACD that humanises service users and emphasises that they have a life outside of their diagnosis, to help reduce power imbalances between mental health service providers and Black service users. Different levels of training where Black service users, carers/supporters, and MHS learned together, so that everyone received the same information and learned from one another, was suggested as a means of addressing power imbalances. The importance of MHS acknowledging the role they or their colleagues may play in the mistrust Black service users have in the mental healthcare system was also highlighted, and that understanding a service users background and cultural experience may increase Black service users trust with services. This involves explicit acknowledgement of historical and individual negative experiences and listening to the experiences that underline the treatment preferences that people express for ACD inclusion. The workshops also reinforced the need for an independent facilitator as they were cited as having the potential to empower Black service users in relation to their care. Furthermore, their involvement was seen to reduce the extent to which ACD creation adds to the existing pressures of MHS including workforce shortages, insufficient role delineation, and resource constraints.

### Summary of the co-production and ACD creation strategies

A steering committee, a lived experience advisory group, and a staff advisory group provided guidance throughout the project. Co-production workshops based on stakeholder recommendations shaped the implementation resource: an ACD template, training, and a manual and job description for the ACD facilitator (23). Two training courses were developed: a simulation training course and a Recovery College course, both for service users, staff, and carers/supporters. The ACD facilitator, a social worker employed by the NHS Trust, was given the manual and created the ACDs of participating service users with their care team whilst undergoing fortnightly supervision. Throughout the ACD creation period, monthly staff meetings were held for staff in participating teams to discuss the project and presentations to clinical teams and third-sector organisations were also delivered. Awareness videos about ACDs were also created and distributed. Throughout the project the resource was improved based on data collected through participant interviews and staff meetings. The strategies not detailed in the methods section can be found in Additional File 1: Additional project strategies. A Recovery College course and simulation training were developed instead of modalities requiring less resources as during this project an e-learning resource was already being developed (28) by colleagues who were being funded by another source, Health Education England.

### Co-production workshops

Based on the recommendations from stakeholders, three co-production workshops were conducted with mixed stakeholder groups to develop: an ACD template; the Trust’s Recovery College course to provide information on ACDs; and skills training delivered with the Trust’s Simulation training team on how stakeholders could create and use an ACD. The development of the ACD template involved seeking views on existing documents (Crisis PaCK (29), Crisis Plus (30), Joint Crisis Plan (17), and MAPS (31)). The trainings workshops sought to determine what people thought was needed to make them effective for all stakeholders. The simulation training workshop allowed the Simulation team to create a bespoke course to include service users and carers/supporters, as they had not previously run a course for these stakeholders.

### ACD facilitator manual and supervision

A co-Principal Investigator (co-PI) (CH) revised a manual drafted after the CRIMSON trial of Joint Crisis Plans (17) for clinicians taking the role of the independent facilitator leading ACD creation. The facilitator was independent in that they were not a part of a service user’s care team. A facilitator was recruited from within the NHS Trust and attended the simulation training and fortnightly supervision via hour long virtual meetings with a project co-PI (CH) who is a psychiatrist. The facilitator was White/of European descent. We did not attempt to match the facilitator to the service users by race/ethnicity, this is because it will not be feasible to do so when ACDs are rolled-out nationally and it is important that all staff are trained to deliver culturally appropriate care (32). Furthermore, this type of matching ignores the diversity that exists within racial/ethnic groups (32).

### ACD creation

The facilitator used the AdStAC ACD template during meetings to create ACDs. Service users were invited to bring any carers/informal supporters. Meetings could be conducted via video call, phone call, or in-person. The initial model was a minimum of two meetings, based on previous research (17), the first including the participant’s care-coordinator (a MHS member who co-ordinates a service user’s care across the health and care system) and the second meeting including their treating psychiatrist; both to support the service user and provide their professional insights. Meetings were scheduled by the facilitator and the research assistant. Mental health capacity was assessed by the MHS member present and the facilitator during each meeting. At the end of the creation of the ACD a confirmation of mental capacity to complete the ACD was recorded on the ACD by the MHS member, which included an assessment of capacity to complete the ACD and an assessment of capacity to make advance choices about refusing treatments. This was done to confirm the refusals and requests made in the ACD are reflective of the service user’s preferences when well and that their capacity to do so was confirmed by an MHS involved in their care.

### Training – Recovery College course

The Recovery College course was developed to provide an opportunity for stakeholders (Black service users, carers/supporters, and MHS) to learn about ACDs. Its development was informed by stakeholder workshops co-led by a peer trainer employed by the Recovery College. The final course was produced through collaboration between the peer trainer and the AdStAC team (facilitator, research associate, and research assistant) and delivered by the peer trainer and facilitator with assistance from the research associate and the research assistant. The peer trainer was Black/of African descent and the facilitator who was a co-trainer was White/of European descent. It was a four-day in-person course, running weekly from 10am-3pm. The course included didactic and interactive components with regular space for discussion and individual and group exercises. Roleplay exercises were not included in the course and a fictionalised example of a Black service user was utilised to explore possible considerations when creating an ACD. Whilst the course was developed to be of use to all stakeholders, the primary target of the course was service users as they make up the majority of Recovery College students.

### Training – Simulation training

The simulation training was developed for stakeholders (Black service users, carers/supporters, and MHS) to develop skills in completing and using ACDs in their existing role. Simulation training replaces or amplifies real experiences with guided experiences that evoke or replicate aspects of the real world in a fully interactive manner (33). It was a full-day of in-person training comprising didactic content delivered by a co-PI (CH) with a barrister (ARK) present to address legal questions; simulated scenarios that simulated ACD completion and use in practice that were run by the training staff (who are clinicians trained in simulation training) with a professional actor (provided with detailed instructions for each scenario) playing the service user; and adapted Pendleton’s debrief sessions facilitated by the training staff. Debriefing followed the format: what went well; what could have been done differently; and the golden moment - a significant point where what was said/not said/done changed the direction of the scenario. The training was run twice to allow modifications based on observation and participant feedback. As mentioned previously, the simulation training was adapted for service users and carers/supporters to take part in addition to the clinicians usually targeted; the training was also adapted as a way to train facilitators and other staff who may be involved in creating or using an ACD.

### Recommended strategies

Table 1 shows the AdStAC activities that map onto the strategies to “enhance the adoption, implementation and sustainability of a clinical program or practice’ and their clusters as defined by the Expert Recommendations for Implementation Change (ERIC) study (34). See Additional file 2: Known barriers and recommended strategies for how identified barriers to ACD implementation characterised by the Consolidated Framework for Implementation Research (CFIR) (35) are addressed by the project strategies.

**Table 1.**
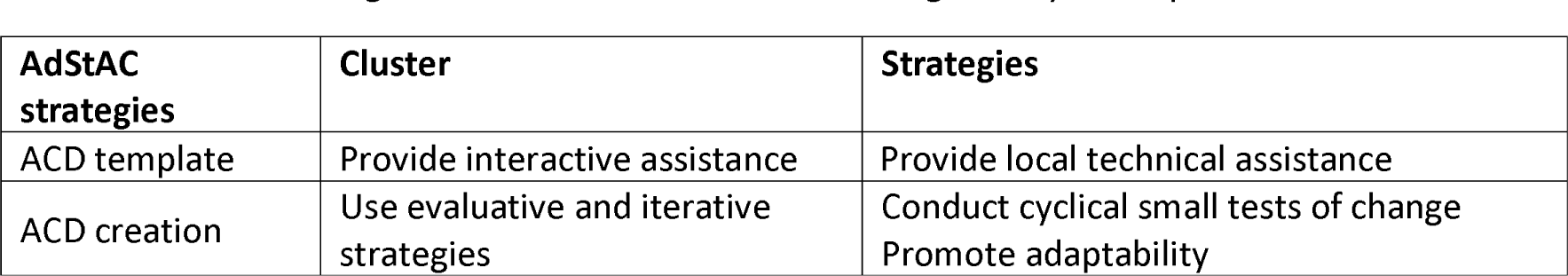
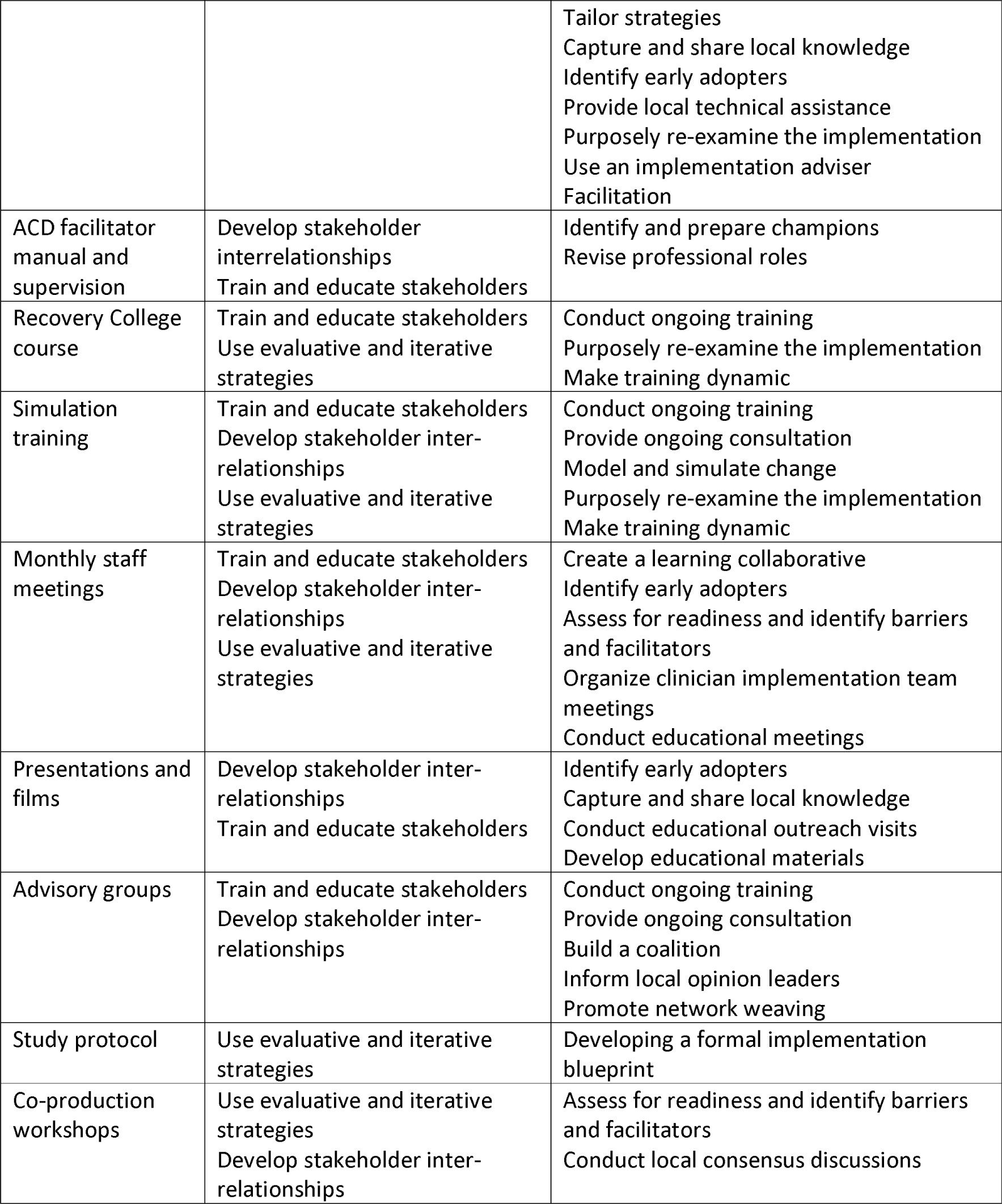
AdStAC strategies and the ERIC clusters and strategies they correspond with.

## Measures, data collection, and analysis

### Recovery College course

Participants were invited to a group feedback session at the end of the course to share their opinions of the content and the experience. The researchers took notes of their responses.

### Simulation training

Participants were asked to complete pre-course and post-course questionnaires rating their knowledge, confidence, and understanding of ACDs. Paired samples t-tests were conducted to compare pre- and post-course answers. Thematic analysis (36) was conducted by a Simulation team member (SGa) on the open-text survey given, which covered people’s experience, what they learned, and, for staff, perceived influence on their practice.

### ACD creation

The Acceptability of Intervention Measure (AIM), Intervention Appropriateness Measure (IAM), and the Feasibility of Intervention Measure (FIM) were adapted for use by service users and staff following the authors’ instructions (37).

Service users’ trust in mental health services was assessed using an adapted single item question: ‘Generally you can trust mental health staff and services’, used previously in research with this population (38) with responses on a five-point Likert scale.

Service users completed baseline and post ACD completion questionnaires including the item on trust in services. Post ACD completion interviews of service users and staff covered acceptability, appropriateness, and feasibility of the ACDs they were involved in creating using AIM, IAM and FIM and open-ended questions about their experience and motivation to complete an ACD. Interviews were recorded and transcribed verbatim for thematic analysis.

The ACD facilitator logged the profession and role of meeting attendees, the number and duration of meetings, and meeting locations. Descriptive statistics were calculated for the lengths of ACD creation meetings, their quantity, who attended, and their location. ACD content was coded for refusals and requests regarding treatment preference when in contact with mental health services during relapse of illness or a crisis.

### ACD creation interviews

All ACD creation interviews were conducted by JS. Interview transcripts were then analysed using thematic analysis (36). This entailed transcripts being coded using NVivo14 and initial themes and a coding framework being developed. The transcripts were then revisited to further develop the themes and coding framework. Once this was completed ABa checked the coding framework against the transcripts to see whether they felt there needed to be any additions or amendments to codes or theme names. JS and ABa are both Black/of African descent. The wider research team also commented on the themes during discussion on quotations to use for the paper.

## Results

### Co-production workshops: resource development

The ACD template workshop led to the addition of a ‘Who I am’ section where people could include content important to their sense of identity that they wanted others to know. It was felt that this type of information could humanise service users and lead to less stigmatisation and more respectful treatment during a relapse or crisis. Later feedback from various sources enabled the template to be modified to include questions around advocacy, rephrasing of its wording to be more sensitive to people’s individual experiences, and flow of its presentation.

The ACD template included the sections personal details (including main difficulties, or considerations, etc.); who I am; care in crisis (including key signs that I am in crisis, helpful things when I am in crisis, key things people misunderstand about me, etc.); preferences for particular services and instances (e.g. community/home treatment team care, Hospital care, Mental Health medical treatments, etc.); Practical help (care arrangements for dependent(s)/pet(s), whom to inform about admission, etc.); how different people can access the document (e.g. ambulance services, mental health staff, general hospital, etc.); confirmation of mental capacity to complete the ACD; summary of views on the ACD of people involved in creating the ACD; and a record of the people involved in creating the ACD, their role, and the date of their final review of the ACD.

The Simulation team determined stakeholders wanted unified learning objectives across roles and then drafted learning objectives. For example, participants expressed the importance of learners understanding the document was patient-led and not like previous documents, like care plans, so one learning objective became “implement a model of decision making that is truly shared between service users, carers, and healthcare professionals when making ACDs”. The team also identified beneficial and acceptable content for simulation scenarios. For example, cultural sensitivity and service user individuality were two desired themes, so the ACD creation scenarios included specific religious and dietary requests, and the scenario actor was briefed to become less distressed if participants were attentive to related content in the ACD.

The Recovery College course was determined to require four sessions to cover learning objectives important for stakeholders.

### Training - Simulation training

Two separate trainings occurred; the second was adapted to allow a later start time to facilitate access by some participants, for example those on sedative medication or those who had care/support commitments. Police input was sought for the development of training, but this was not obtained during the project.

#### Demographics

The simulation training was attended by 12 trainees in total, nine recorded as female, three as male, most were over 55 years old (n=5), and the majority identified as of a Black ethnic group (n=9). The professions recorded were nursing (n=1); medical (n=1); allied health professional (social worker[n=1]); other clinical professional (peer support [n=1]); other non-clinical professional (carer [n=2], senior management [n=1], peer advocate [n=1], volunteer visitor [n=1]). Two people did not answer the profession question. Whether someone was a service user was not recorded as this was not data routinely collected by the Simulation team, which hitherto had only delivered courses for professionals. No trainees from the simulation training took part in the Recovery College course.

#### Quantitative findings

Paired sample t-tests were conducted on the responses to pre- and post-training questionnaires, see Table 2 for results. Higher scores denote increase in people’s rated confidence, understanding, and ability. Six trainees were excluded for providing incomplete responses for their learning outcomes in the evaluation survey, the resulting data was insufficient for inclusion into the analysis.

**Table 2.**
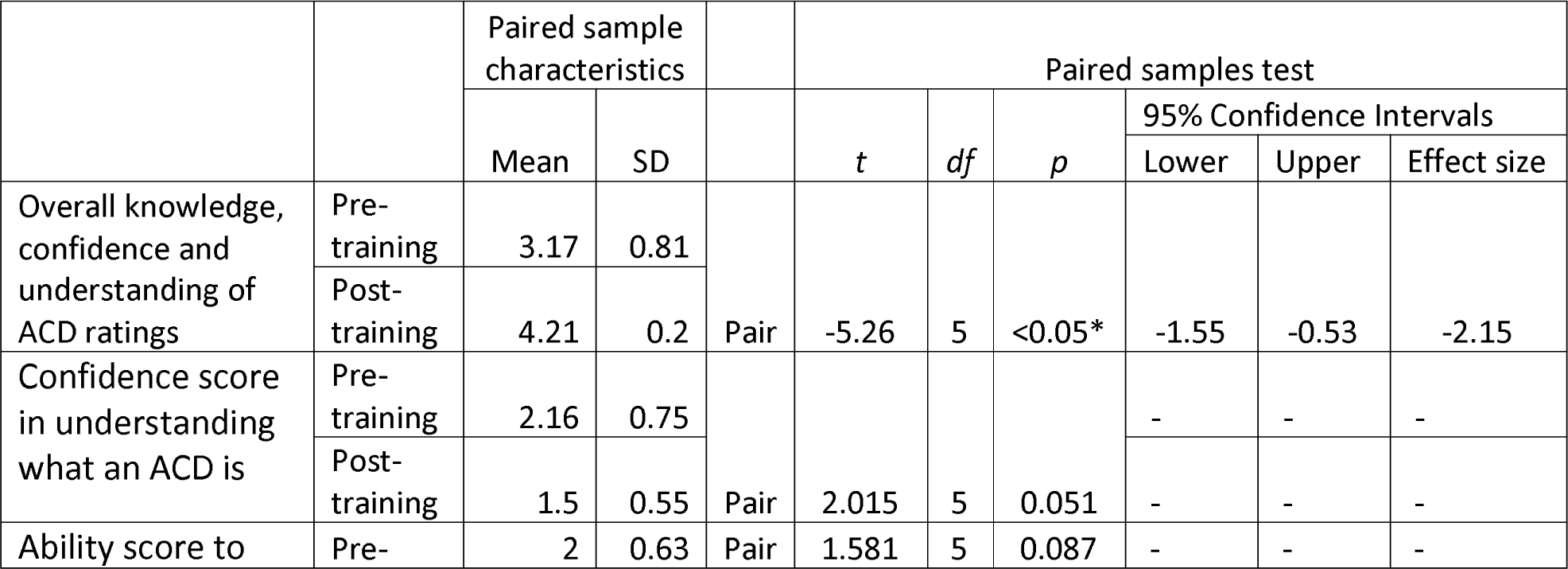
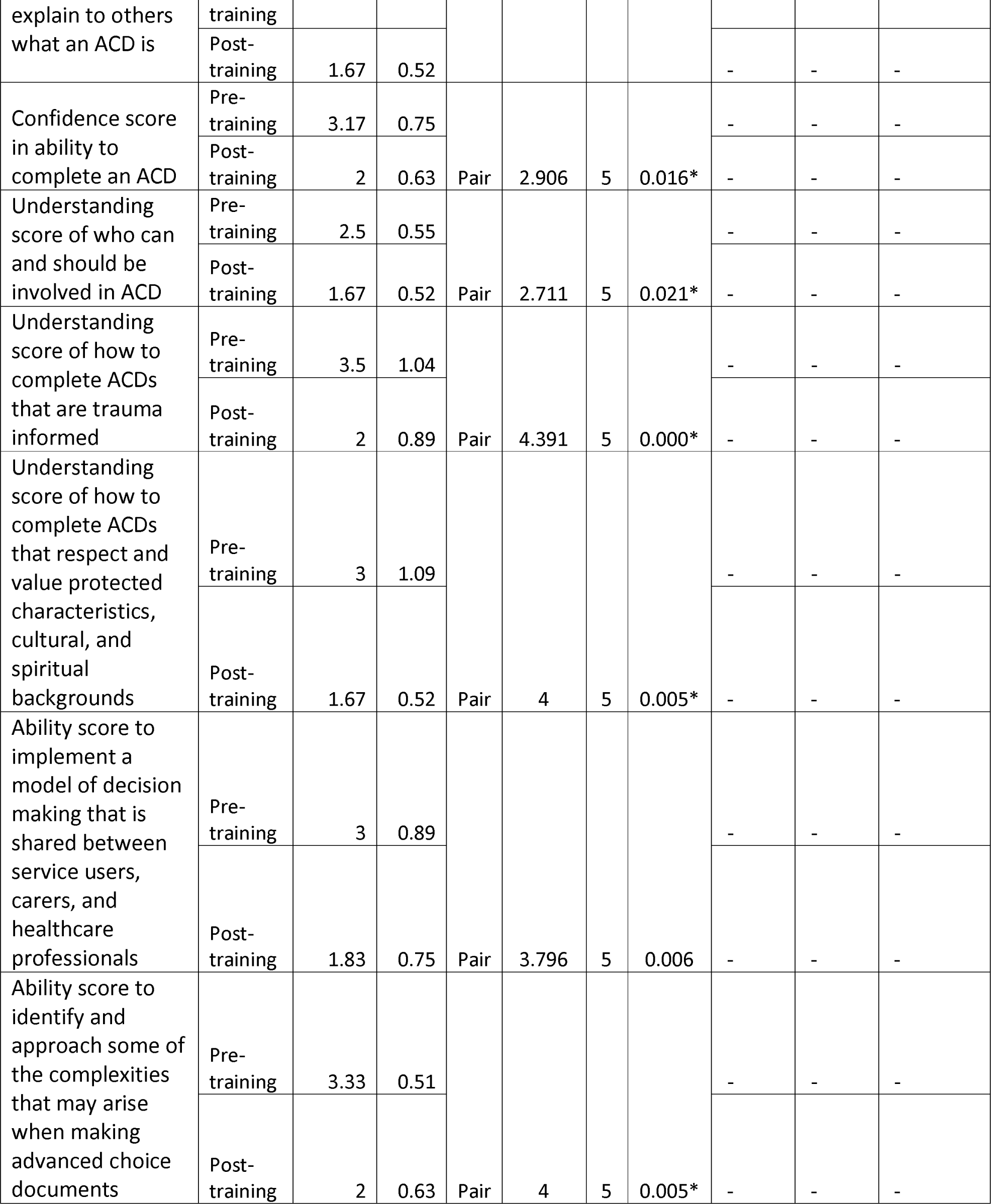
T-test results of pre- and post- simulation training survey scores.

#### Qualitative findings

Four themes were identified through thematic analysis, including increased knowledge from facilitators and speakers; helpfulness of the activities; learning the importance of teamwork and collaboration; and desire for further exploration within the course (including medication refusal, simulation exercises, and how ACDs will work in practice).

### Training - Recovery College course

#### Demographics

The Recovery College course was attended by 12 trainees, ten recorded as female, two recorded as male, with a mean age of 52 (range: 41-62yrs). The ethnicities of the trainees were not recorded as this information is not routinely collected by the Recovery College. Trainees included people currently accessing mental health services (n=5); carers/supporters (n=3); third-sector organisation staff (n=2), and peer supporters and lived experience advisors & experts (n=2). No mental healthcare professionals attended. Six people attended all sessions, four people attended three sessions, one attended two sessions, and one attended one session. No trainees from the Recovery College course took part in the simulation training.

#### Feedback

Trainees described the course as informative and appreciated it being co-facilitated by a peer trainer who detailed their own views and experiences, as it made them relatable and created a safe space where trainees felt free to express themselves. Another favoured element was how interactive the sessions were. However, trainees stated the ACD and what it relates to makes it a ‘heavy’ document, which made the course feel somewhat challenging and so there was a need for a more digestible approach.

Generally, trainees felt more knowledgeable about how to approach making their own ACD with the appropriate people, such as MHS. Additionally, they felt they could support someone else with the making of their ACD. Trainees also felt the course would benefit friends and family of service users; peer support workers; voluntary organisation staff; carers; and mental and general health staff.

Trainees remarked that four days of training was a big commitment, as was the session length (10am-3pm). Views were mixed concerning clinical staff attendance: some thought there should be the option of a group including staff, due to the importance of having stakeholders learn from one another, whilst others did not want this as they did not feel it would be possible to create as supportive and safe a space with staff present.

### ACD creation meetings

See Table 3 for demographics of service user participants. The race/ethnicity of MHS that took part in meetings was not recorded. No service user participants that created ACDs attended the Recovery College course or the simulation training before or after the creation meetings. Nine ACDs were made through an average of two meetings, lasting for an average of 62 minutes. Of the 20 meetings held, 11 were attended by care-coordinators, four by consultants, four by psychologists, one by a case management officer, and one by a team leader. Six meetings took place in-person, and 12 by video call. Six ACDs were not completed due to lack of responsiveness from care teams to setting up meetings or providing input to ACDs through alternative methods offered; care team members not attending agreed meetings; numerous care team changes, facilitator responsiveness, and service user disengagement. Typically, it was simpler to arrange meetings with psychologists, followed by care-coordinators, then consultant psychiatrists, with the latter being more likely to attend once given the option of attending for 15 minutes instead of the full hour. Approaches to scheduling were adjusted with the aim of reducing delay before meetings were held and the ACD template adjusted to maximise information gathering.

**Table 3.** Demographics of service user participants.

| Category | Total (n) |
| --- | --- |
| Age |  |
| 16-25 | 1 |
| 26-35 | 5 |
| 36-45 | 4 |
| 46-55 | 4 |
| 56-65 | 1 |
| **Recorded sex** |  |
| Female | 9 |
| Male | 6 |
| **Ethnicity** |  |
| Black African | 6 |
| Black British | 5 |
| Mixed | 2 (Afro-Caribbean & white English and Scottish; Black Caribbean and white Irish) |
| Black Other | 1 (Black British Caribbean) |
| Black Caribbean | 1 |
| **Highest level of formal education** |  |
| University undergraduate degree | 6 |
| Secondary school education | 4 |
| Post secondary education | 4 |
| University postgraduate degree | 1 |
| **Housing situation** |  |
| Own house, flat or apartment (including council) | 12 |
| Live with others (in their house) | 3 |
| **Living arrangement** |  |
| I live alone | 8 |
| My parent(s) or relatives | 4 |
| My children | 3 |
| **Employment status** |  |
| I'm not able to work (disabled) | 6 |
| I'm looking for a job | 3 |
| I work part-time | 3 |
| I work full-time | 2 |
| I work as a volunteer (not paid) | 1 |
| **Reported Diagnosis** |  |
| Schizophrenia | 3 |
| Paranoid schizophrenia | 3 |
| Psychosis | 3 |
| Bipolar disorder | 2 |
| Schizoaffective disorder | 3 |
| Anxiety and depression | 1 |
| EUPD | 1 |
| **Agreement with diagnosis** |  |
| I agree fully | 8 |
| I disagree fully | 3 |
| I agree partially | 3 |
| Not sure | 1 |
| **Age of first contact with services** |  |
| 16-25 | 9 |
| 26-35 | 3 |
| 36-45 | 2 |
| 46-55 | 1 |
| **Number of admissions** |  |
| 1 | 1 |
| 2-5 | 9 |
| 6-10 | 3 |
| More than 10 | 2 |
| **Number of admissions under section** |  |
| One | 3 |
| Some | 4 |
| All | 8 |

The most frequent request made in ACDs were support (e.g., support groups, cultural support); whom to notify as contact persons; strategies for de-escalation and reduction of coercive measures; additional information and well-being factors (e.g., being outdoors, creative activities, support to take medication); request for other treatment (e.g., psychotherapy); request for medication form; refusal of specific medication; and request for hospital alternative (e.g., home treatment, outpatient treatment) (Table 4).

**Table 4.**
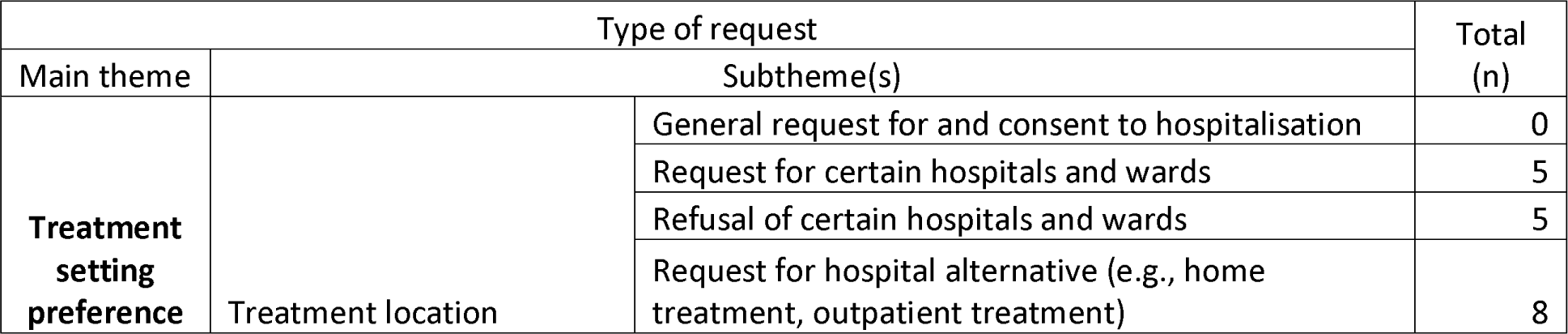
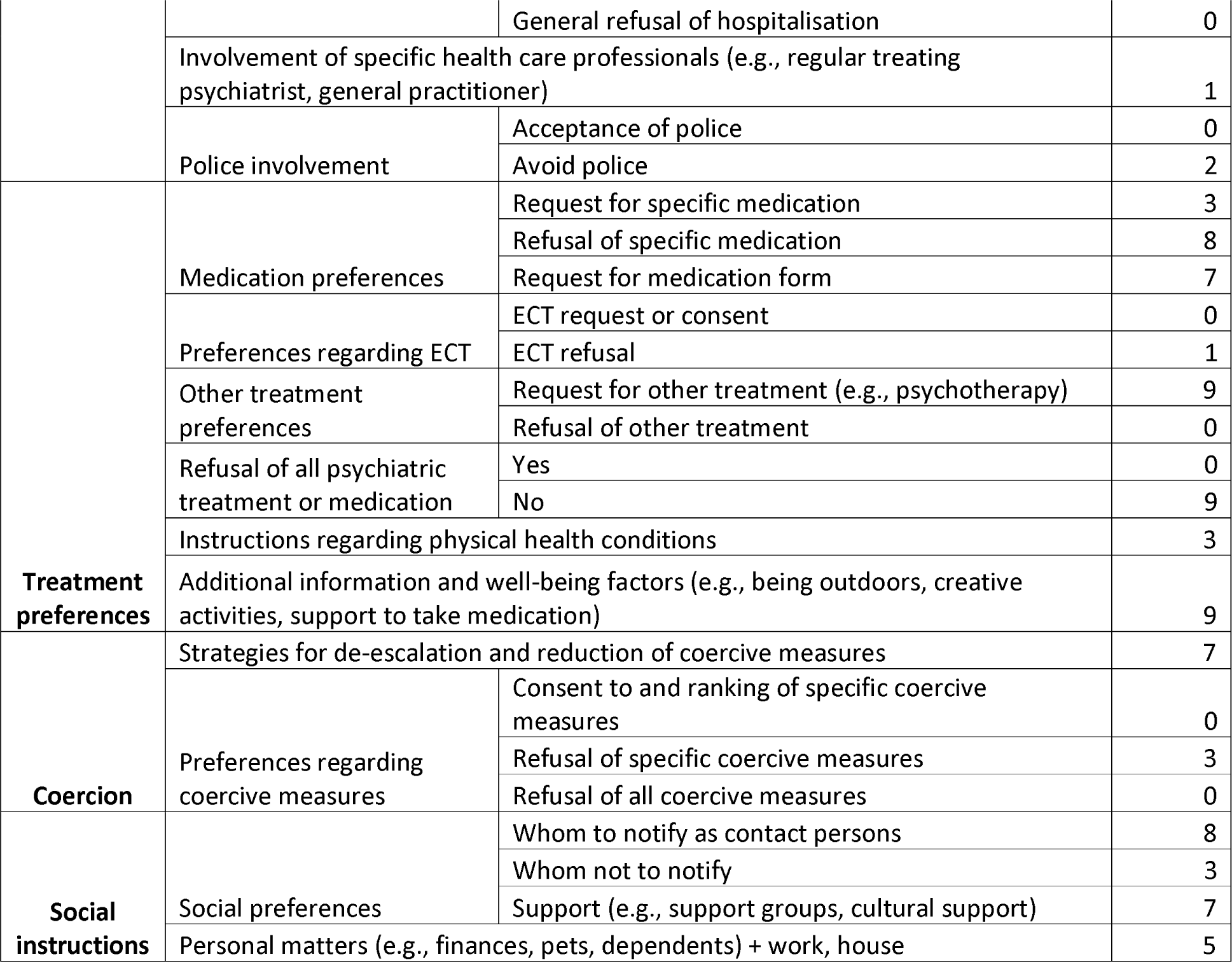
ACD Requests and refusals.

Completed ACDs were stored in the participant’s electronic medical records as a PDF file, with an alert on the face page of the record stating ‘X has made an Advance Choice Document (collaborative advance directive). Stored under correspondence date xxxx. STAFF MUST READ if: they present in crisis, a MHA Assessment is being undertaken, they are admitted to a ward, inpatient care planning is required. It contains Advance Statements valid under [Mental Capacity Act] and MHA Code of Practice guidelines.’

### ACD creation interviews

#### Demographics

Seven service users and five staff took part in the post ACD completion interviews. Two service user participants who had also completed their ACDs did not take part in these interviews, as one was unwell, and one declined to take part.

#### Trust, acceptability, appropriateness, and feasibility

At baseline, service users responded most frequently with ‘Agree’ when asked whether they can trust services. At post-completion ‘Agree’ and ‘Neither agree nor disagree’ were the most frequent responses, with one respondent changing from ‘Disagree’ to ‘Neither agree nor disagree’ (see Table 5). For service users, ‘Agree’ was the most common response across acceptability, appropriateness, and feasibility for their ACDs. ‘Completely agree’ was the second most frequent for acceptability and feasibility had the greatest number of ‘Neither agree nor disagree’ answers (see Additional file 3: Service user ratings of ACDs). For staff, across appropriateness and acceptability ‘Completely agree’ was the most common response. ‘Agree’ was most frequent for feasibility, which elicited the only neither ‘Neither agree nor disagree’ response (see Additional file 4: Staff ratings of ACDs).

**Table 5.** Service user ratings of their trust of services at three study points.

|  | Baseline | Post-completion | Second Follow-up |
| --- | --- | --- | --- |
| SU1 | Strongly agree | Strongly agree | N/A |
| SU2 | Agree | Agree | N/A |
| SU3 | Agree | Agree | N/A |
| SU4 | Agree | Agree | N/A |
| SU5 | Disagree | Neither agree/disagree | N/A |
| SU6 | Neither agree/disagree | Neither agree/disagree | N/A |
| SU7 | Neither agree/disagree | Neither agree/disagree | Neither agree/disagree |
| SU8 | Disagree | N/A | N/A |
| SU9 | Strongly agree | N/A | N/A |

#### Service user interviews

See Table 6 for service user interview excerpts, and Additional file 6: Service user interview excerpts for those not listed here.

**Table 6:**
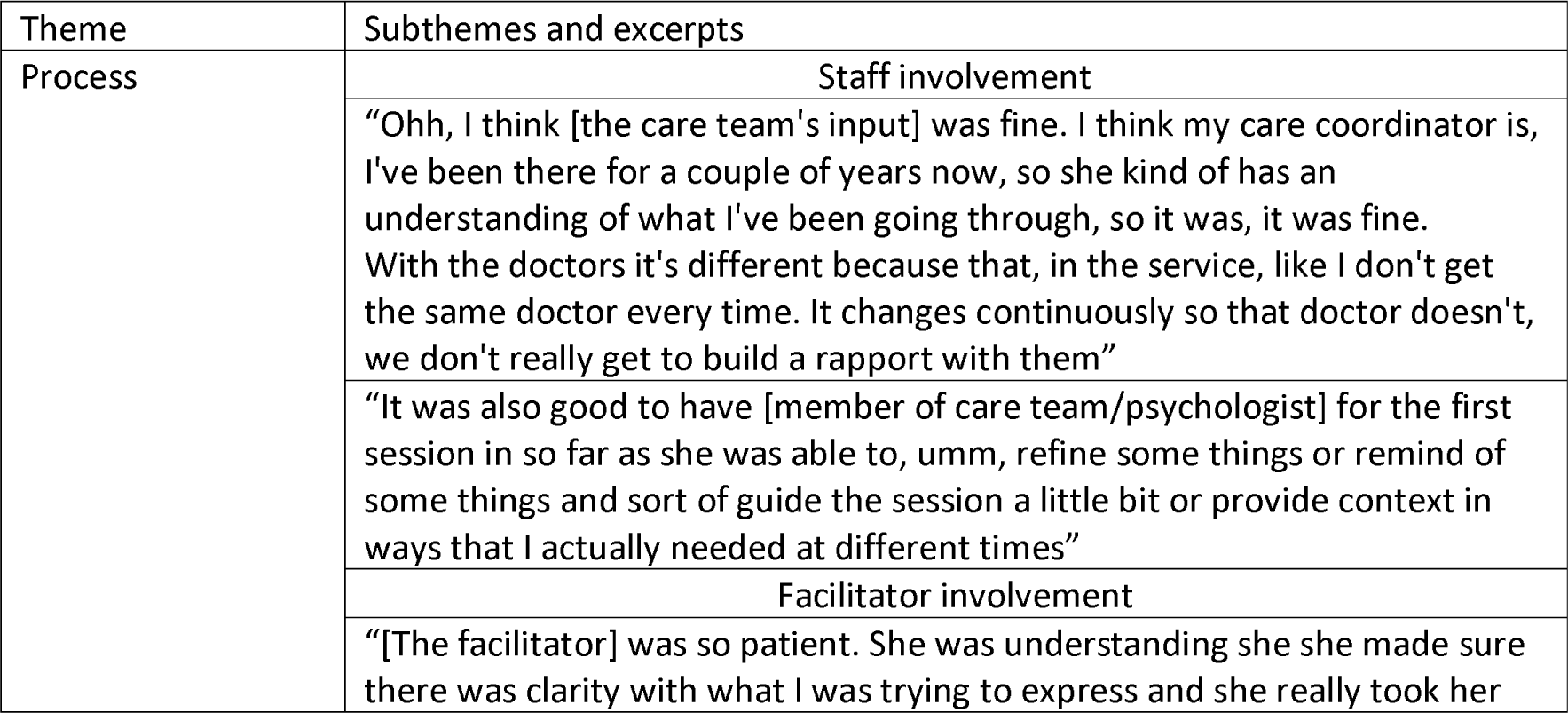
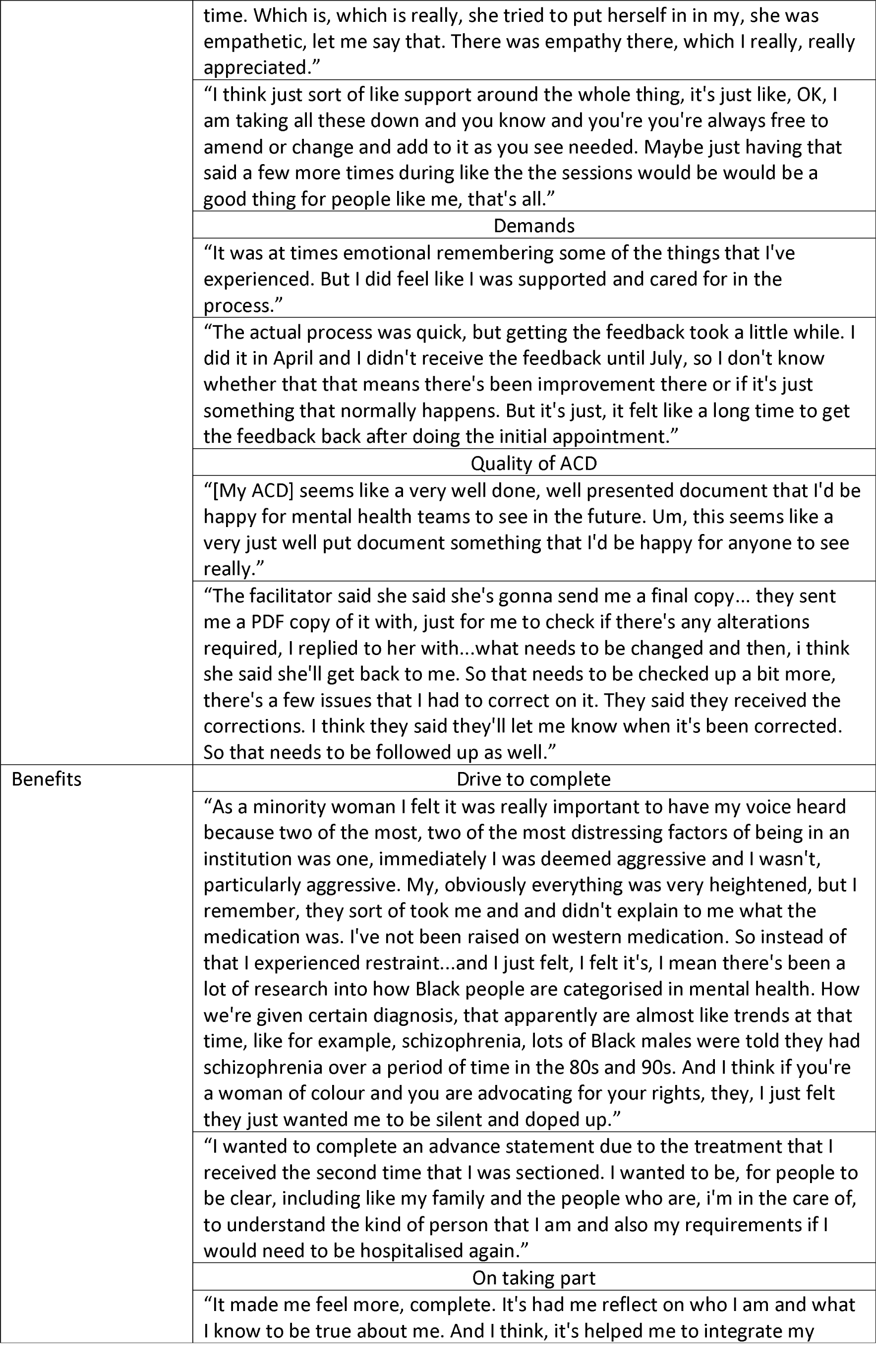
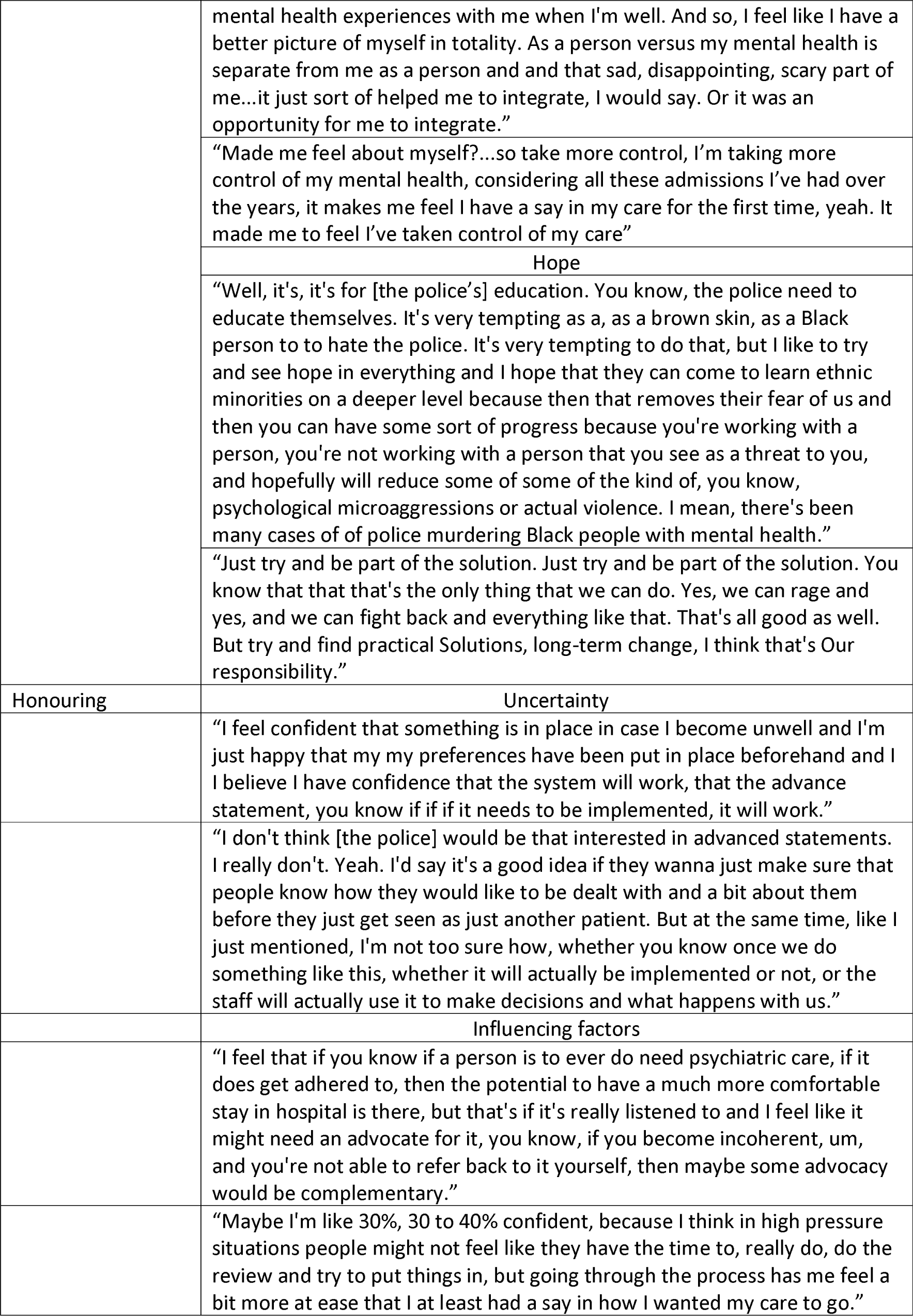
Service user interview excerpts.

##### Process

###### Facilitator involvement

Generally, service users appreciated the presence of the facilitator. They cited their prompts as helpful, although service user SU6 stated that being reminded more explicitly that the ACD was a living document more often would be good. The presence and guidance of the facilitator was also seen as supportive and caring. The ease of someone else recording and summarising was also highlighted as a positive.

###### Staff involvement

Opinions on involvement of a service user’s care team varied. Some saw it as helpful as they were able to provide important insight, and some cited their familiar presence as helping them participate in the creation meetings. Others had a neutral opinion, or noted that attending staff had no input in the meetings, without expressing any opinion on this. The brevity of staff attendance and their absence in some cases were mentioned but no explicit evaluation of this was given.

###### Demands

Some interviewees described the meetings as emotionally demanding and the cause of unease; SU3 stated it can be a daunting task. No participants reported feeling pressured in meetings when asked; however, SU7 didn’t feel able to challenge the facilitator’s summary when asked to review an initial draft of their ACD. The length of meetings was seen as appropriate for the content of the meetings and not demanding, but the time between meetings and between the last meeting and receiving ACDs to review was described as long.

###### Quality of ACD

Service users described their ACD as well made, comprehensive, and a good summary of who they were as a person and their experience. However, SU2 stressed that the ACD was only an indicator of who they were. Most asked for corrections to be made but most did not express an issue with what had to be adjusted. However, SU7 expressed that they felt their racialised experience of services, an experience important to them, was left out. After edits to their ACD were made in response to this, SU7 felt satisfied with the final ACD.

##### Benefits

###### Drive to complete

Participants cited multiple reasons for choosing to make an ACD, including being heard and increasing their autonomy. A desire to increase understanding was also mentioned multiple times, primarily in staff, but also family. Related to this, some people also wanted to avoid the repetition of coercive treatment and negative experiences with mental health services.

###### On taking part

Service users reported positive feelings for themselves and towards services. For services, participants detailed having a confidence in them, feeling listened to and respected, and being appreciative of having the opportunity to express themselves. SU3 however had an unchanged opinion of services, as they reported previously having confidence in them. Participants expressed finding the experience empowering and having a greater sense of control and feeling more involved in their care. Some found the experience itself enjoyable, for SU1 this was because they were being heard. Others were pleased with themselves for taking part and shared that it was a way to appreciate themselves due to the reflection involved, and others felt they had increased in self-confidence. The ACD creation process was also appreciated for its personalised approach, which was described as humanising by some. SU7 also mentioned how the ACD provided reassurance for carers/supporters, which was of value to them.

###### Hope

Participants shared that they were hopeful creating the ACD would bring about better treatment for themselves when in contact with mental health services and be a part of a more holistic approach to their care. SU7 also described the potential of ACDs to reduce police aggression and violence against people from minoritised groups.

When asked what they would say to someone considering ACDs, participants cited the support or facilitation of support for their mental health needs as a reason to complete it. Additional reasons were its ability to increase understanding of who they are as an individual and to have their care personalised. Related to this, participants also mentioned having more say in their care and empowering themselves. The ACD was also spoken of in terms of safety and it being a safety net, with SU7 describing it as a practical solution to racial inequalities.

##### Honouring

###### Uncertainty

Uncertainty was widely expressed about whether ACDs would be used. Service users stated either they could not know until it was in practice or that they weren’t familiar enough with ACDs to say. Some said their satisfaction with creating an ACD was dependent upon how it would be adhered to/honoured. Others were sceptical about whether it would be used, with some citing having low confidence in services or police using it, due to previous experiences. Participants also expressed uncertainty when asked what they would say to people considering ACDs, because they themselves were sceptical or inexperienced with ACDs. However, SU4 and SU1 had a general confidence it would be used.

###### Influencing factors

Participants felt the honouring of their ACDs depended upon staff training and awareness of ACDs. Furthermore, participants felt how staff and police regarded ACDs would have an impact on whether they were utilised. SU7 felt that for ACDs to be honoured an advocate would need to be involved. SU1 believed their own behaviour would have an influence on whether people would adhere to their ACD. Participants also generally felt that staff capacity and services in general being overwhelmed presented a risk of their ACD not being honoured.

###### Staff interviews

See Table 7 for staff interview excerpts, and Additional file 7: Service user interview excerpts for those not listed here.

**Table 7:** Staff interview excerpts.

| Theme | Subthemes and excerpts |
| --- | --- |
| Process | Approach |
|  | "I thought was a really nice collaborative process. I know that the service user we did it for, certainly enjoyed it. It was really beneficial having...an external facilitator kind of come in and do that as well, I think because it just guided our thinking. So yeah, it was, it was, I thought, was really helpful, as did, as did my client." |
|  | "There was a bit of a delay, which is, you know, probably inevitable in the paperwork being completed, erm which might have perhaps meant things went off the boil a little bit. Umm, I've actually still got some sections to look at err to complete, yeah....so I think I think between us having the interviews and the actual advance statement being written up for my clients comments and mine and the doctors erm you know there was several months. So, I think maybe you perhaps lose a little bit of momentum...Umm yeah, but I'm, I'm not, that's not a criticism, you know, I understand people are, you know, overwhelmed." |
|  | Care team involvement |
|  | "There was one point when I erm where I kind of stepped in and wanted to kind of highlight and, you know, what, how somebody might misunderstand here. What might somebody see and how might that not be your normal self? Because it felt like compared to what I knew of the client and what they've shared with me in CBT, they were being a little bit more cautious in terms of what they were stating for the document." |
|  | "The facilitator didn't know the client, but I was present and I did, so I was able to do that prompting and that might be the case for most people who complete it, that just the the fact of having somebody there who knows that means there's that bit of prompting if somebody isn't being that assertive...this was a client who was particularly self-critical, so [they] might say kind of well if I was coming across as kind of angry or aggressive or blah blah blah would describe things in quite judgmental language and I was kind of prompting [them] to be like, what exactly might you be saying? How else might somebody experience that erm rather than to kind of be critical of [themselves] in [their] describing of what [they] might be like when [they] was well, unwell, sorry. And so yeah, it might just be a facilitator is able to kind of keep an eye out for when people's self-esteem or self-criticism might impact on the completion process and and be ready to give a bit of a prompt if someone is using quite kind of, yeah, self, self-disparaging or self-critical language." |
|  | Independent facilitator impact |
|  | "One of the advantages I noticed that patient were able to talk about things they haven't been mentioning before in a consultation and with their meeting with with a care-coordinator, but they might be able to, there was, the patient was able to talk about it in, while we're being completing them, advance statement, and I think it give us a bit, little bit better understanding when patient might stop or refuse a treatment and a kind of, when we understand why it's happening, it's a field we can work on to just to improve kind of compliance and overall well-being." |
|  | Yeah. So, in terms of the experience of getting them completed, it was a really positive experience erm with the client that I referred. I think she found it really useful and it was really helpful to kind of hear how kind of unbiased and open all of the questioning was with with her. So rather than |
|  | trying to lead her to a particular response or erm entry in the document, it felt like there was lots and lots of effort to make sure it really reflected her preferences and her wishes. Erm, it seemed really, really helpful. |
| Benefits | For staff |
|  | I haven't used [an ACD] yet, but I do know it's quite helpful. I mean my background is inpatient and community as well and I do know when people come in in a really terrible state, they don't, they can't tell you a lot, so I think knowing what their wish would be in terms of treatment helps a lot, professional, help professionals a lot to kind of make sure we're doing the right thing in supporting that person in that, in their recovery. |
|  | "I do a lot of CBT for people with bipolar disorder, so advanced planning and relapse prevention planning is part and parcel of that work all the time erm but certainly this project has prompted me to think a bit about how the person's kind of colour and cultural background might have a particular impact on their experience of mental health services in those crisis points." |
|  | For service users |
|  | "I think it's very useful to have our, umm, Black service users making advance statement. As we all know, there is this unconscious bias about Black service users using the service and the treatment they've had in the past. So, it's it's helpful that we can get their voice in the treatment and and also, yeah, and it gives them the confidence of accessing the service too, because a lot of them do not access the service for the simple reason that they know a lot of people in the past has not had the, had had been treated, I would say perhaps differently, but I know that's all changing as well. So it's nice to give them that confidence that the service is changing and they they've been heard and them having an advanced statement will be helpful in their recovery as well" |
|  | "So, I felt it was a very therapeutic interview. We did two, I I was involved in two actually erm I thought it was very positive about kind of, you know, uh, capturing my client's kind of personality that things that were important to [them] and the some of the distressing experiences [they'd] had. So, it did feel very positive." |
| Application | Barriers |
|  | "I think erm you know, it would be important to have some kind of meetings with with relevant staff so that it is really given a high profile, because otherwise it may not got followed and then that would be detrimental actually if people have gone to the bother of doing it and then feel it's not being followed. That would be, you know, sort of worse actually than not doing it. You see what I mean? So that slightly concerns me because all these things are emergencies. I happen to be an [Approved Mental Health Professional]" |
|  | "I think for other people that I've used advance statements with where their preferences might be quite strong in terms of 'I'm always offered this medication, I hate this medication, I really don't want that, I want something different.' I can imagine that there might be more challenges with implementing it, because somebody might read well it's your preference, but that's not what I think you should have, and there might be more of a tension between what a team thinks is the right thing and what the person has requested." |
|  | Facilitators |
|  | "I think some something maybe where there was kind of champions or |
|  | whatever, so there was somebody who would, you know, in the ward who would be responsible for, you know, yeah erm disseminating the information. Erm so that it, you know, like you might have a lead on the ward for carers or something. Somebody who would take on that role on on wards erm, because otherwise communication is notoriously difficult erm, because it's essential on the wards that this information is there." |
|  | "To my recollection, [they] didn't request anything that would be really, really outside of normal care practices. So rather than thinking do, you know, actually people have a tendency to really misunderstand me and they really need to kind of pay lots of attention to this document before I receive any treatment otherwise, things can go terribly wrong. It felt more like some of [their] things were about I'd slightly preferred this over this and if I am on a ward, these are the kind of things I would like these are people that need to be contacted. So I think it'd be quite feasible for it to be followed." |

##### Process

###### Approach

Staff remarked how the process of creating the ACD felt collaborative in a way that felt personalized to the service user. Staff also approved of the ease of the process and the flexibility in respect to scheduling and meeting location. Whilst participants said no improvements were needed, it was mentioned how the delay in meetings and getting completed ACDs back was an issue that could impact on service user motivation.

###### Care team involvement

Some participants highlighted the need for a member of the care team to be present in meetings as their insight can be uniquely useful; particularly if a service user is being reserved, or alternatively an independent facilitator would need to be vigilant about this. Additionally, staff noted that a service user’s wellness was a factor that needed to be monitored and that their care team was well suited to advise on this. Some staff shared that they did not feel an independent facilitator was essential for the ACD to be created; MHS6 felt it was unlikely they’ll be available when ACDs are upscaled, so the process needs to be adjusted for care teams to facilitate ACD creation.

###### Independent facilitator impact

Staff shared how the presence of the facilitator as an independent party allowed for a safe space to be created, which was supportive for both staff and service users. In this space it was said that there was an increased sense of openness that led to conversations that were not had in regular interactions. The involvement of a neutral party was also seen as beneficial in reducing the impact of staff conflict of interest, who may have their own priorities regarding a service user’s care that differ to the service users. Staff frequently commented on the positive input of the facilitator and their sensitivity in dealing with challenging topics and their making the process a personalised experience.

##### Benefits

###### For staff

Generally, participants saw the ACD as informative and a useful tool, with MHS3 saying that it could provide targets where it was not previously known what works for service users or there was a lack of understanding about an aspect of their life. MHS4 also shared how the project being focused on Black people and then creating the ACD with that in mind made them think about how someone’s colour or culture could impact their experience of mental health services, in a way they hadn’t considered before in their work. The process itself was also said to be rewarding for staff, with MHS1 finding it enjoyable.

###### For service users

Staff saw the ACDs as an opportunity for service users to: be heard and have more input in their care; to prepare for their future care; preserve their rights during their contact with services; and to be given a sense of safety that they will be looked after in their preferred way. ACDs were seen as part of a holistic approach to care through more personalised action, and MHS3 noted they saw ACDs as a continuation of community care. Staff mentioned the importance of ACDs being completed by Black service users due to social inequities and hoped that having ACDs would provide them the confidence to access services when needed. MHS5 raised that due to the sensitive work involved in creating an ACD and the hope that it can offer, not honouring an ACD would have a significantly negative impact on service users.

##### Application

###### Barriers

Staff reported they would reserve stating how confident they were in ACDs until they were used in practice. They mentioned that whether ACDs would be honoured would be dependent on both requests and context, for example, which team was involved and in which clinical setting. Communication on wards particularly was highlighted as a barrier, as was collaboration between teams. MHS3 cited the latter as requiring proactiveness with ACDs. Staff thought that it would be easier to communicate the need to check an ACD with community mental health teams than with ward teams, and MHS5 felt confident the community teams would access it. Staff were unsure how police would interact with ACDs, due to not having much contact with them, but mentioned how police are busy and react to things quickly and that this would likely affect whether they would check whether someone has an ACD or adhere to it. MHS4 felt that if a service user had recurrent contact with police, they may be prompted to access their ACD.

###### Enablers

Staff awareness of a service user having an ACD or knowing what they were and how to use them was seen as key to them being honoured. ACD champions were one solution suggested to ensure clinical teams were made aware and accessed them. Participants also expressed concerns about how easily they could be accessed. They wondered what their visibility would be like on the electronic clinical records system used and highlighted the importance of them not being lost in patient notes. Staff also stressed the importance of the ACD being connected with legislation as an important part in garnering interest and faith in ACDs for service users. Generally, a wider rollout was sought to realise the potential benefits of ACDs. ACD completion when service users were well was underlined as crucial for achieving the benefits of ACDs for service users.

## Discussion

The project set out to develop implementation resources to get ACDs to work for Black people and those that support them. An ACD template, two sets of training, and a facilitation process were developed and used in the creation of ACDs and the development of awareness and skills associated with ACDs and their use. In addition to gaining insights into the meaning of ACDs for service users and staff, the project identified key issues for the upscaling and routine use of ACDs.

Training of staff may not be achievable through the developed courses. The Recovery College course, which was developed into a four-day course, had no clinical staff attendance, likely due to the time commitment required and lack of an official requirement to attend. This absence does not fulfil the joint learning requested by stakeholders in workshops; however, the trainees largely accepted the lack of staff attending and some preferred this. Simulation training trainees generally reported finding the training useful for practical application in various stages of creating and using ACDs. A noteworthy finding was that they felt less confident to complete ACDs than before they started. This could be due to them believing they knew what ACDs were beforehand and then finding that they were different to what they expected, and perhaps more complex. The reach of this training’s observed value may be limited as simulation training needs to be delivered in small groups.

The completion of the ACD creation meetings requires a degree of swiftness from staff to maintain momentum and to avoid fluctuation in a service user’s mental state that results in them being deemed to not have capacity to create an ACD, which can be hard to achieve. Service users expressed how demanding the process could be; and staff believed it was important to time the meetings for when service users were well and to complete ACDs promptly to avoid demotivation. However, staff cited high workload and higher priorities as reasons for not attending or rearranging meetings, which increased time spent arranging meetings.

As there were substantial delays in setting up first or second meetings, due to lack of staff availability or response, some meetings went ahead without staff present. It was found that the presence of a care-coordinator was not always necessary for productive meetings; however, one instance showed that having someone who knew the service user at the first meeting was crucial in establishing the context of discussions. Additionally, although staff presence was sometimes not considered vital by service users, some noted that it was reassuring.

Staff generally approved of the process of creating ACDs and described the creation and use of ACDs as well aligned to community care practice. This contrasts previous findings of resistance to the role of the facilitator, doubts about ability to follow preferences, and beliefs that the process duplicates existing practice (20). However, it does support findings that some MHS believe having an independent facilitator can lead to the successful completion of ACDs, particularly due to their impartiality and having time to complete the document (39). Although some staff felt that a service user’s care team could facilitate ACD creation, this was partly due to scepticism regarding the use of facilitators being scaled-up rather than resistance to their role.

Though service users generally reported positive views about the meetings, the act of creating an ACD, and their ACD, echoing previous research findings (17,29,40), people from historically marginalised groups and those who have individual negative experiences with services may still find it difficult to speak freely when invited to by MHS. Discussion of racialised experience is not routine, as exemplified by the staff member who required prompting to consider how a service user’s lived reality as a Black person may influence their experience of mental health care, and the service user who had an additional meeting to discuss following encouragement from JS during an interview. JS’s being Black/of African descent may have generally encouraged service user participants to discuss their racialised experiences to the extent that they did, and at various points during the project participants did mention JS’s race/ethnicity as a favourable part of the project. This project may also have partly facilitated this, through the project title and its self-identified race/ethnicity-based eligibility criterion, but project observations suggests that more explicit encouragement is needed. This may cause discomfort in meetings already found to be challenging by service users; we therefore suggest that this encouragement is made in initial offers to make an ACD, so that service users can consider discussing these experiences beforehand, and that staff and facilitator training covers the need to discuss these experiences as part of ACD creation.

The content of service user ACDs varied from including personally known people in their mental health care, reducing staff coercion and how to do this, the way they received their medication, what medication they did not want, specific requests for psychotherapy or art therapy, letting MHS know what additional things help them, such as particular creative activities or being outside, and requests for treatment at home when possible. Such requests if honoured have the potential to lead to the strengthening of therapeutic relationships, fewer hospital admissions, and an increased sense of safety and being supported in a personal manner. It is noteworthy that there were no refusals of all coercive measures, but none consented to or ranked any particular measures. Also, a minority mentioned the police in their ACD, despite either having past interactions with police or recognising the possibility of their involvement in the future.

Both service users and staff reserved judgement on the likelihood of ACD content being honoured, with staff describing awareness, content, and service context as influencing this. Staff capacity was also cited as an issue in ACDs being honoured, due to staff overwhelm and the NHS being under pressure overall. Advocates and staff ACD champions were suggested to help with staff accessing and honouring ACDs in busy environments such as wards. Champions on wards are generally accepted as playing a significant role in implementation (41,42) and culturally appropriate advocates have been said to be useful in having service users’ needs met and rights adhered to (43). These points highlight how ultimately for ACDs to work for Black service users, in addition to specific strategies, a culture prioritising the service user’s voice is essential. This further highlights the need for ACDs to have legislative backing and having considered reasoning for not honouring them. Scepticism also existed around whether police would read or honour ACDs irrespective of training, due to them not seeing it as priority. As the ACDs made as part of this project were only accessible via medical records, and police cannot access these, participants were not asked whether they wanted to specifically permit or refuse police accessing their ACDs.

### Strengths and limitations

Carrying out this study within one service provider allowed the creation of a resource that is feasible for use in its context and was largely acceptable to participating service users and staff. The focus on Black service users also enabled the development of resources that aim to improve mental health equity, resources well suited for further improvements and adaptions with this aim in mind. The project therefore established a useful foundation to apply and evaluate these resources on a larger scale. The ACD creation model developed in this project also presents a means to adhere to the supported decision mandate of the United Nations’ Convention on the Rights of Persons with Disabilities (44). This is due to its collaborative, service user led nature that supports people with long-term mental health difficulties to exercise their right to refuse and request treatment, particularly in instances where they may be deemed to not have the capacity to make such decisions. However, due to the context specific nature of the study, its applicability to other mental health services provider contexts may be limited; for example, while Recovery Colleges are widespread (27,45), access to simulation training may vary. The close involvement of researchers throughout the project also means that the extent to which the results reflect use of the implementation resource in typical practice is limited. The small sample size also limits the findings’ generalisability, and the lack of staff attendance at the trainings limits what can be said about the usefulness for the everyday practice of staff and whether this varies between staff types. No forensic service users or staff were included in the study despite attempts to do so and given the disproportionate number of Black people in such services (46), the resources need to be piloted and amended as required for use within these services. Although carers/supporters were involved in the co-production phase and attended both sets of training, none were included in ACD creation meetings. Further study is needed to find out why and how better to include them. The confidence in the findings from the AIM, IAM, and FIM implementation outcome measures is also limited, due to the repetitive nature of the questions. Importantly, the time frame of this project did not allow follow up to ascertain rates of access to and honouring of ACDs, nor to develop and test a process for their review. The Advance Choice Documents Implementation project (47) which is run by our group and is based on AdStAC seeks to expand on the findings of AdStAC, in part by addressing such limitations as detailed here.

### Implications

Adaptations are likely to be needed to optimise implementability in other services and in other populations. In addition to the need to adapt and pilot ACDs in forensic services, this will also be needed for child and adolescent mental health services and those for older adults.

Further implementation studies will need to consider both additional strategies and research questions. Besides training that can easily be accessed by staff, other strategies are likely to be needed to increase the agility of community staff to respond to requests for ACDs. The digitisation of ACDs is likely to aid efficiencies in ACD creation and development, quality, safety monitoring, and routine clinical notes retrieval, and so should also be considered an important part of ACD implementation. Strategies are also needed to enable discussions about care influenced by other characteristics such as gender identity, sexuality, religion, etc. One possibility is that the role of the independent facilitator be expanded to promote this change in culture. Likewise, strategies will be needed to encourage relevant professionals to follow the content. While we did not encounter the same attitudinal barriers that have affected previous research, we cannot assume they are now non-existent. Furthermore, it still needs to be determined whether trial findings regarding the use of the MHA and therapeutic alliance can be replicated through routine use, even with implementation resources and strategies in place, for example due to trial samples not being representative of wider populations. This includes the question of whether ACDs can improve mental health equity. These questions need to be approached with observational and quasi-experimental study designs. Finally, there is no evidence about predictors of whether ACD content is accessed and followed, including characteristics of services, mental health staff, service users, or ACD content. This question requires large sample sizes, follow up lengths sufficient for periods when ACDs should be accessed, and informatics work. Future work by our group will address these questions to fully realise the potential of ACDs in addressing inequity in mental health care and enhancing the impact of mental health services more widely.

## Supporting information

Additional file 1 Additional strategies

Additional file 2 Summary of CFIR barrier constructs and the related ERIC and AdStAC strategies

Additional file 3 Service user ratings of the acceptability, appropriateness, and feasibility of their ACD

Additional file 4 Staff ratings of the acceptability, appropriateness, and feasibility of ACDs they worked on

## Data Availability

ACD resources are available on request from CH at
Quantitative data are available on reasonable request; however, qualitative data and ACD content are not due to risk of reidentification.

## Abbreviations

AdStAC: Advance Statements for Black African and Caribbean people project
ACD: Advance Choice Document
AIM: Acceptability of Intervention Measure
CFIR: Consolidated Framework for Implementation Research
co-PI: co-principal investigator
ERIC: Expert Recommendations for Implementation Change
FIM: Feasibility of Intervention Measure
IAM: Intervention Appropriateness Measure
MHA: Mental health act
MHA review: 2018 Independent Review of the UK Mental Health Act
MHS: Mental health staff
NHS: National Health Service

## Additional files

File name: Additional file 1

File format: Microsoft Word Document, DOC

Title of data: Additional project strategies

Description: Project strategies not directly a part of the implementation resource described in the paper

File name: Additional file 2

File format: Microsoft Excel Worksheet, XLS

Title of data: Summary of CFIR barrier constructs and the related ERIC and AdStAC strategies

Description: Summaries of the CFIR constructs (barriers) to ACD implementations matched with ERIC strategies to implement ACDs and the AdStAC strategies that correspond

File name: Additional file 3

File format: Microsoft Excel Worksheet, XLS

Title of data: Service user ratings of the acceptability, appropriateness, and feasibility of their ACD

Description: Service user participant responses to AIM, IAM, and FIM measures after ACD completion

File name: Additional file 4

File format: Microsoft Excel Worksheet, XLS

Title of data: Staff ratings of the acceptability, appropriateness, and feasibility of ACDs they worked on

Description: Staff participant responses to AIM, IAM, and FIM measures after ACD completion

## Declarations

### Ethics approval and consent to participate

Approval for the study was granted by the Bradford Leeds NHS Health Research Authority Research Ethics Committee (REC reference number: 22/YH/0012) on 07/02/2022. All study participants provided informed consent before they took part in the study.

### Consent for publication

No identifiable information of individuals included in the manuscript.

### Availability of data and materials

ACD resources are available on request from CH at Quantitative data are available on reasonable request; however, qualitative data and ACD content are not due to risk of reidentification.

### Authors’ contributions

Advisory groups – AS chaired the staff advisory group meetings. SGi chaired the lived-experience advisory group meetings.

ACD template workshop – ABa, CH, and SS facilitated the co-production workshops.

Simulation workshops and training – ABa, CH, MF and MS facilitated the co-production workshops. ABi, AKS, CH, MF and MS facilitated the training, with support from ABa and JS. SGa analysed data collected from trainee surveys.

Recovery College course workshop and training – ABa and PE facilitated the co-production workshops. FC, PE, ABa, and JS developed the course. FC and PE led the facilitation of the course, ABa and JS supported. ABa and JS held and recorded the feedback session.

ACD creation – ABa, CH, and SS developed and revised the ACD template, JS also revised. CH developed the facilitator manual and led facilitator supervision. FC facilitated the creation of service user ACDs. JS collected baseline and follow-up data, transcribed interviews and analysed transcripts. ABa checked the created coding framework for reliability. FH led monthly staff meetings, supported by CH and JS.

Films – SS, SGi, and JS were a part of filmed interviews on ACDs.

Manuscript – JS led on drafting and editing the manuscript. All other authors reviewed, edited, and approved the final manuscript.

## Acknowledgement

Within the NHS Trust - Recovery College; Maudsley Learning; Patient and Carer Race Equality Framework team; Staff BME forum; Lewisham Crisis Plus team; Simone Myers, Lewisham EDI lead; and Southwark: Trauma Informed Care Implementation Group. Staff and lived experience advisory groups; AdStAC project steering committee; Black Thrive: Culturally Appropriate Advocacy and Peer Support service; and Croydon BME forum.

