## Additional file 1 Additional strategies for "Developing and testing Advance Choice Document implementation resources for Black African and Caribbean people with experience of compulsory psychiatric admission"

### Additional project strategies

##### Advisory groups – Steering committee, Staff, and Lived Experience

The steering committee was chaired by the Chair of the Board for the NHS Trust. Other members comprised: clinicians with experience of Advance Statement legislation in Scotland; the chair of the Trust Patient and Carer Race Equality Framework; the Trust Clinical lead for digitisation; a Professor of Mental Health and Inclusivity; a barrister whose expertise is in human rights, particularly in relation to vulnerable people; and the Chief Executive of a local forum for Black and Ethnic Minority people. They advised on delivery and dissemination.

The Staff advisory group was chaired by a study co-applicant (AS) who is a professor in mental health nursing. The group comprised staff who work across acute and community mental health services and were identified as opinion leaders, with the ability to influence colleagues to adopt the intervention and advise on staff engagement. The lived experience advisory group was chaired by a study co-applicant (SG) who is a serious mental illness lived experience and anti-racism consultant who identifies as biracial. This group comprised Black service users with a previous experience of compulsory admission and carers/supporters of Black service users. Questions for the workshops were developed, results of stakeholder workshops and recommendations on the procedures and materials for implementation were discussed with each group. The lived experience group advised on recruitment and participation of service users.

##### Monthly staff meetings

Monthly information exchange meetings were held virtually for staff in participating clinical teams or those interested in the project. These meetings were hosted by the ACD facilitator supported by a co-PI and the research assistant. Their format consisted of a presentation recapping the project and ACDs, followed by discussion.

The monthly staff meetings were largely attended by staff who were interested in learning more about the project. As a result, the sessions mainly comprised information giving to staff on ACDs and the process of referring people to the project and subsequent steps. Staff recommended ways to encourage service users to participate (where to promote, types of meetings, particular contacts), team procedures (e.g., how to book appointments with consultants), how to make ACDs routine practice (e.g., promote as helpful for care plans). Common questions and concerns included access to ACDs, legal standings of ACDs (e.g., the implications if a team overrides or a service user wants something different), being spread thin but wanting to be involved and conflict resolution (discrepancies between clinical judgement and service user desires). Also, staff felt it would be hard to get service users near discharge to participate if they had difficult past experiences with services. Attendees expressed being in favour of ACDs, hoping that it would be cathartic for service users and help reduce how negatively they feel about services, in addition to reducing misunderstanding around service user’s preferences and cultural considerations.

##### Presentations and films

In addition to presentations made to the groups summarised in the project strategies, two filmed interviews with members of the AdStAC team were made. One comprised a co-PI (SS) who is a consultant psychiatrist and another consultant psychiatrist who was identified as an opinion leader. This interview was tailored to a clinical audience and covered the background to ACDs and addressed common queries about their use. The second was an interview with the chair of the lived experience advisory group by the research assistant, which covered the benefits of ACDs and the experience of creating one. Both interviews were uploaded to an online video player and the link to this upload shared via presentations, emails, the Trust’s internal and external website, and social media. Additionally, the interviews were uploaded as part of a Health Education England funded web resource.
